# Effect of implementing digital adherence technologies on the use of health care providers’ time and the human resource cost of tuberculosis treatment adherence support in Ethiopia

**DOI:** 10.1101/2024.03.14.24304331

**Authors:** Mahilet Belachew, Mamush Sahile, Demekech Gadissa, Tofik Abdurhman, Ahmed Bedru, Christopher Finn McQuaid, Job van Rest, Kristian van Kalmthout, Degu Jerene, Katherine L Fielding, Amare W Tadesse, Nicola Foster

## Abstract

**Background:** The management of human resources for health is a key priority for resource constrained health systems. Digital adherence technologies (DATs) for treatment support offer an opportunity to reorganise existing healthcare worker workload. Historically, tuberculosis treatment services involved directly observed treatment with frequent patient visits to the clinic for treatment adherence support. More recently, digital technologies have offered an option for the digital monitoring of treatment adherence and remote support. The aim of this study was to determine how health care workers use their time following the implementation of digital adherence support, and the human resource cost of tuberculosis treatment adherence support.

**Methods:** As part of the ASCENT three-arm cluster randomised trial of DATs for tuberculosis treatment support, we conducted time-and-motion studies in 15 of 78 ASCENT-trial health facilities for six-weeks. Healthcare workers recorded time spent on direct patient care and in-person adherence monitoring for 10 patients, as well as time spent on above-service activities. Total time use was calculated by multiplying time spent per visit by number of patient visits at the facility for six weeks and adding above-service time. In a facility-level analysis, we used negative binomial regression models to investigate associations between total time use and facility-level factors.

**Findings:** HCWs spent 4% (126/2892) of time in minutes on in-person adherence monitoring in the Labels, compared to 7% (206/3126) in the Pillbox and 13% (587/4673) in the standard-of-care study arms. More time was spent on above-service activities in the standard-of-care (2360 minutes [SD:1951]) compared to the DAT arms (Pillbox: 2057min [SD:1570]; Labels: 2261min [SD:1360]). After adjusting for facility workload and number of people lost-to-follow-up, we did not find sufficient evidence of a difference in the human resource costs between the study arms (Labels IRR (95%CI): 0.71 (0.33; 1.52); Pillbox (95% CI) 0.71 (0.33; 1.52)).

**Conclusion:** We found a small, although not statistically significant reduction, in human resource costs in the facilities where DATs were implemented. We observed changes in how healthcare workers spent their time, including a shift towards fewer health facility visits but more time spent on in-person adherence support. Further work is ongoing to assess the implications for the cost-effectiveness of the interventions.

**Author summary:** Digital adherence technologies (DATs) can be used to support patients taking tuberculosis treatment by monitoring how often and when they take their medication. Healthcare workers can then access this record electronically and provide additional support to patients if it is found that they are not taking their medication. We compared the tasks that health care workers do between health facilities using digital adherence technologies and those without. We found that patients made fewer visits to health care facilities using DATs than the standard of care - but when they did attend, they spent more time per visit speaking with health care workers than those without DATs. Healthcare workers similarly spent more time on adherence support in the facilities with DATs but less time on tuberculosis care overall because of each patient having fewer visits.

## Introduction

The World Health Organization (WHO) global strategy on human resources for health 2030 is aimed at improving development outcomes by ensuring adequate investments for health services [WHO 2016] [1]. Achieving these goals is constrained by the limited availability of healthcare workers in many countries, including Ethiopia. In 2010, Ethiopia had an estimated physician-to-population ratio of 1:42,706 and 1:5,000 for nurses and health extension workers [2]. Historically, tuberculosis treatment has been a human resource intensive activity because of the Directly Observed Therapy (DOTS) treatment strategy that requires supervised medication taking [3]. Digital Adherence Technologies (DATs) such as smart pillboxes (Pillbox) and labels (an SMS-style intervention) provide support for the monitoring component of the treatment strategy, allowing patients to take their treatment at home. As an indirect measure of treatment adherence, through DAT engagement, their dose-taking is logged with each pillbox opening, and reviewed by healthcare workers in real time, negating the need for frequent and costly visits to the health facility. During the implementation of a new technology, we expect adaptation of routine practice as the new technology gets assimilated [4]. How the technology gets used in routine practice will influence whether the innovation improves patient outcomes and reduces healthcare worker time spent (efficiency). There is currently a lack of empirical data describing how the implementation of DATs affects the practice and workflow of healthcare workers in routine settings.

Currently, typical workload activities at the tuberculosis clinic include providing tuberculosis treatment, managing adherence and adverse drug reactions, TB and HIV collaborative activities, referring extra-pulmonary tuberculosis and drug-resistant tuberculosis patients to higher level facilities; support health facility staff in non-tuberculosis clinical care; keep patient records and manage stocks of medicines and other supplies; and planning and implementing TB and Leprosy control [5]. These activities are typically performed by one or two health professions per facility. The Ministry of Health (MoH) of Ethiopia is planning to integrate training of tuberculosis management during preservice training. TB management training for healthcare service staff is given post-service and is resource intensive. Currently, TB-delegated HCWs are busy due to a high workload and overlapping activities besides tuberculosis-related duties, such as primary care [6]. Multiple visits for daily DOT adherence support significantly contributes to this workload. Monitoring of TB treatment adherence using digital adherence technologies (DAT) could potentially reduce this workload, as well as help TB patients to take ownership of their treatment journey and overcome TB-related stigma. Implementing a differentiated care approach could improve treatment outcomes, minimizing unnecessary patient health facility visits and costs for both health care system and patients.

This study is part of the ASCENT trial, evaluating the implementation of two DAT interventions in two regions of Ethiopia, Addis Ababa and Oromia [7]. The DAT technologies used are the smart pillbox (evriMED1000) and medication labels (a SMS-style intervention) alongside the DAT platform and differentiated care. It was hypothesised that using the digital adherence technology platform would save health care providers time spent on adherence monitoring and provide clear guidance for follow-on actions required when treatment adherence is sub-optimal. The aim of this study was to compare (i) the time spent by health care providers on activities required to provide tuberculosis treatment and (ii) the cost of supporting adherence monitoring between facilities using Digital Adherence Technologies (DATs) and those providing the standard of care (SoC).

## Methods

### Study design and intervention

The ASCENT trial is a three-arm pragmatic cluster randomised trial, conducted in the Addis Ababa and Oromia regions of Ethiopia. A total 78 health facilities in predominantly urban areas were randomised, in a ratio of 1:1:1, to either Pillbox, Labels or Standard of Care (SoC) study arms. For the digital adherence technologies (DATs) study arms, each dose taken by the patient is logged on the linked web-based data platform automatically either when the pillbox is opened, or when patient texts the hidden code printed on the dose label to a dedicated toll-free number to represent a dose taken. Healthcare workers use the data platform or mobile phone app to evaluate treatment adherence and update support provided including phone calls or home visits to the patient. Further details of the trial and associated health economics work is available from the protocol [7] and health economics analysis plan [8]. To compare human resource time spent and support costs between study arms, we conducted a time-and-motion study as part of the ASCENT-Ethiopia trial between October 2021 and June 2022 in a sample of 15 health facilities (five in each arm) participating in the trial. Facilities were selected based on study arm, and to ensure inclusion of facilities from urban and rural settings. Trial registration: PACTR202008776694999.

### Study procedures

Over a six-week period, data were collected in each of the sampled facilities using two surveys. Facilities completed their evaluation period between October 2021 and May 2022. The first survey recorded healthcare workers’ patient-level interactions per visit, by self-report (survey A). The TB focal person responsible for TB care completed a form at the end of each patient consultation, recording the amount of time they spent on various activities including history taking, diagnostic tests, preparing and dispensing treatment, and care for non-TB conditions. This survey was completed for ten consecutive patient consultations. The second survey (survey B) recorded time spent on tasks when the patient is not in front of the healthcare worker. At the end of each of six weeks, the TB focal person recorded the time spent that week on reviewing adherence records, calling patients on the phone because of missed doses, home visits because of missed doses, and time spent on administrative tasks recording patient details for patient management. For each health facility, data were collected on the staff category of the person providing TB treatment (TB focal person), where the salary costs associated with each category of staff employed in the Ministry of Health were then collected separately. These data were used to estimate the value of staff time spent in each facility. All costs were collected in local currency (Ethiopian Birr) and converted to US Dollars (USD) using the average exchange rate for 2021 to 2023 (48.14 ETB = 1 USD).

As part of the trial procedures, each facility recorded the number of patients attending the clinic using a screening and enrolment log, as well as the number of health facility visits for the entire treatment period of study participants. We restricted our analysis to only facility visits that occurred during the six weeks of the evaluation period, in the 15 facilities sampled. Additionally, staff supporting health facilities to implement the interventions kept logs of visits and calls they made to the health facilities to support the implementation of the technology including training provided. The data for surveys and health facility visit logs were entered onto a REDCap platform.

### Data Analysis

We used an ingredient costing approach to estimate the provider costs of human resources, training and supporting the tuberculosis treatment adherence intervention in the sample of 15 of the 78 health facilities participating in the trial [9]. The time spent by health workers recorded on the time-and-motion studies, were valued using the staff grade and associated salaries of the tuberculosis focal person (one nurse) at each of the health facilities. The number of health facility visits (workload) for each health facility were calculated from the health facility visit log that recorded all tuberculosis treatment related visits of patients enrolled in the trial; and the screening and enrolment log which recorded number of treatment initiation visits made at the health facility. The measurement and valuation of the costs and events included in this analysis is summarised in **Error! Reference source not found.** with additional details on the cost analyses in the appendix (see SS1).

We defined human resource time spent as the time spent on (A) direct patient care activities and (B) time spent on above service activities related to tuberculosis care and adherence support. The additional activities associated with treatment adherence outside of the patient consultation are considered as above service activities and include phone calls, home visits, administrative related activities, and patient adherence record reviews. Time spent on direct patient care was calculated by multiplying the average total time spent on patient-level activities per visit (from survey A) by the total number of patient visits per facility during the six-week period. We estimated this separately based on time spent per patient in the intensive phase and number of intensive phase visits; and time spent per patient in the continuation phase and the number of continuation phase visits. The six weekly observations per facility (survey B) were added together to get a total above-service time spent. Total time spent for the six weeks per facility was calculated by adding the patient-level and facility level time spent together. Time spent per patient was then estimated by dividing total time spent by the number of patients receiving tuberculosis care at the facility during the six-week period. Staff time costs were estimated by multiplying time spent (in minutes) by the salary of the TB focal person in that facility (in cost per minute). In addition to the cost of staff time estimated, we also estimated the cost of training healthcare workers in the use of digital adherence technologies and tuberculosis treatment adherence support guidelines. We also included the cost of implementation support which included visits and support calls made to staff implementing the intervention to health facilities, supporting the use of the technologies.

### Statistical analysis

All data were analysed using SPSS and Stata version 18 In a facility-level analysis, we used negative binomial random effects regression models with the amount of time over which total costs were estimated (6 weeks) as the denominator to model total human resource and investment costs of support between the SoC and Pillbox as well as the SoC and Labels arms of the study. The main outcome used in this analysis was total cost per facility. The variance of cost between study arms was greater than the mean, making negative binomial models more appropriate than Poisson regression models of count data. This was assessed using goodness of fit chi-squared tests. We conducted sensitivity analyses to explore how key analytical decisions affect our estimates of the cost of human resources and support. Firstly, we increased the frequency of retraining from every 6 months to every 4 months and secondly, we increased the cost of healthcare workers’ salaries to represent a scenario where the intervention is being scaled up to include the private sector.

### Ethics

The study was approved by the ethics committees at the London School of Hygiene & Tropical Medicine (approval reference: 19120); World Health Organisation Ethical Review Committee (approval reference: ERC.0003297); the Addis Ababa City Administration Health Bureau Public Emergency and Health Research Directorate Institutional Review Board (approval reference: BEFO/HBTFH/1-16/10415).

## Results

A total of 150 patient-level observations (10 patients each from 15 facilities, survey A), and 90 above-service level observations (6 weekly observations from 15 facilities, survey B) were collected. A summary of the number of health facility visits recorded during the six-week period, by visit type and compared between study arms is reported in Figure 1. SoC facilities had a greater average number of visits (143.1) compared to the Labels (35.4) or Pillbox (42.7) facilities during the six-week evaluation period, despite similar average number of patients seen per facility with 21.4 (SD: 7.6) in SoC, 16.6 (SD: 6.1) in Labels facilities, and 21.4 (SD: 4.4) in Pillbox facilities. In all study arms, there were more frequent patient visits during the intensive phase of treatment than the continuation phase, which aligned with TB treatment guidelines in Ethiopia.

**Figure 1.**
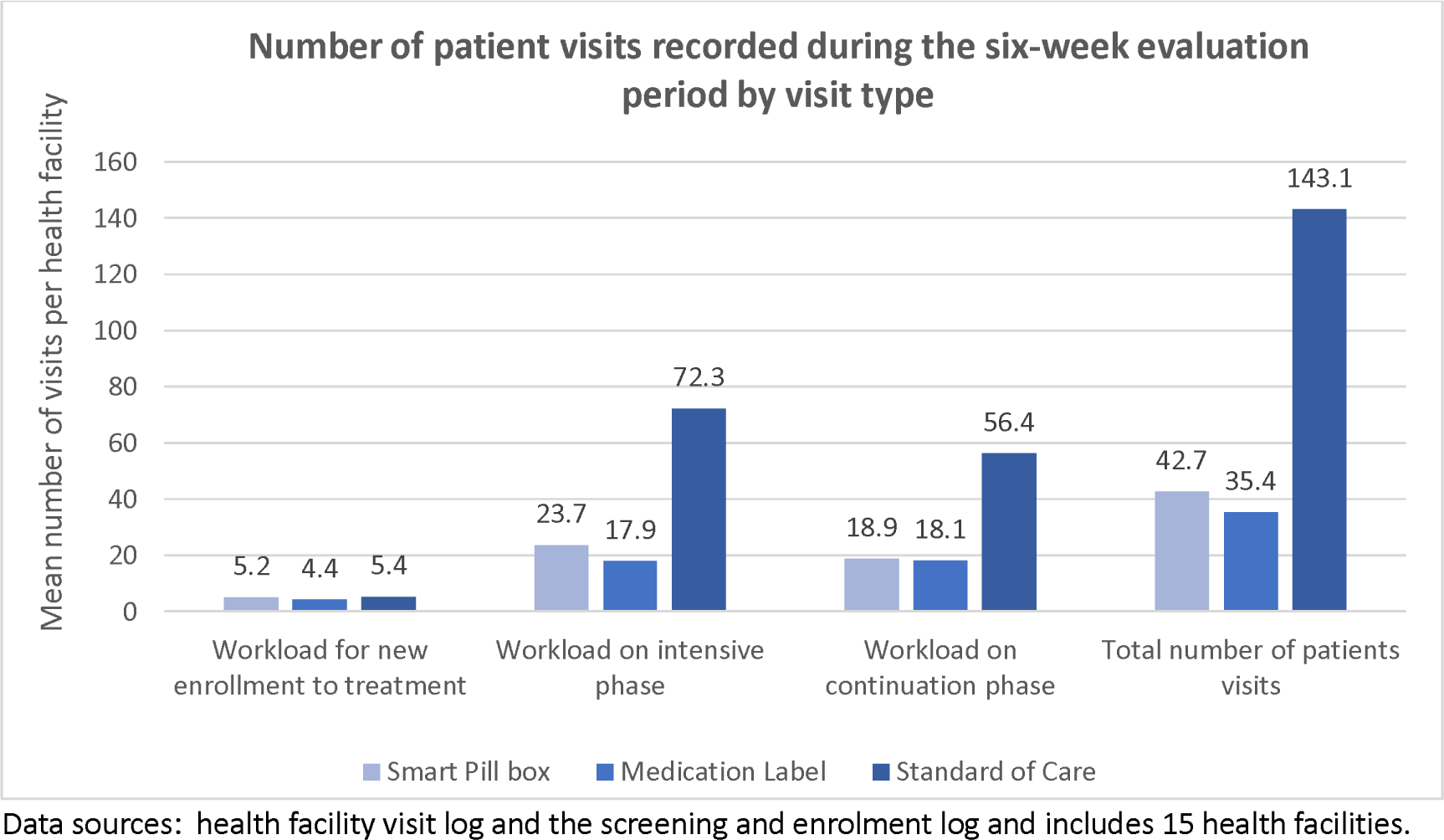
Summary of number of patient visits per health facility by visit type during the six-week evaluation period.

**Table 1.** Summary of the valuation of costs and events included in the analysis.

| Description of indicator | How measured | How valued |
| --- | --- | --- |
| Human resource cost of time spent on direct patient care. | Survey completed by healthcare workers upon patient leaving the consultation. One healthcare worker per facility in 15 health facilities recorded their interactions with 10 consecutive patients over the six-week period. . | Time was valued based on the salary grade of the TB focal person employed at the health facility, and the time spent on such activities was converted to cost. |
| Human resource cost of time spent on above-service activities. | One healthcare worker per facility in 15 health facilities completed a survey at the end of each of six weeks, reporting how much time they had spent on activities such as administration, record-keeping, and adherence support activities. Time was allocated to the per-patient level using an allocation factor based on the average number of patients seen at the health facility during the 6-week evaluation period. | Time was valued based on the salary grade of the TB focal person employed at the health facility, and the time spent on such activities was converted to cost. |
| Number of patient visits to the health facility during the six-week evaluation period | Using a log kept at the health facility, recording the dates of patient visits for patients enrolled into the study. Additionally, used number of first treatment visits recorded on the screening and enrolment log to count the number of patients visiting the health facility who were not enrolled into the trial (this includes patients <18 years, those with extra-pulmonary TB and those who would not stay for the next 1 year in the study area). | Used as the allocation factor for allocating time spent without patients at the health facility down to a patient-level. |
| Training costs | Data collected by staff at KNCV Ethiopia on the cost and frequency of training of health care workers at health facilities | Average salary cost of a nurse or doctor in the Ethiopian health sector. Cost per patient was |
|  | on the intervention. | estimated by allocating the costs incurred across all 78 health facilities to the 15 selected facilities using a health service utilisation allocation factor and to the six-week period of evaluation by including 22.5% (6/27 weeks) of the cost to represent the six-week evaluation period. |
| Support costs | All visits and calls (plus the time taken for each visit/ call), made by KNCV staff to health facilities to provide support in the implementation of digital adherence technologies were logged on the project web-based app. In addition to this, we included 50% of the annual salary cost of a KNCV technical officer supporting the implementation of the technology.<br>KNCV (Royal Dutch Chemical Association) provided implementation support to the health facilities. | Time measured was valued using the salary cost of the TB focal person employed at the health facility where the survey was completed. Cost per patient was estimated by allocating the costs incurred across all 78 health facilities to the 15 selected facilities. This cost was then divided by number of patients seen with 22.2% (6/27 weeks) of the cost allocated to represent the six-week evaluation period. |

When we compare the number of visits per patient during the six-week evaluation period between the three study arms (see Table 2), we find that patients in the SoC arm had an average of 4.7 (SD:2.1) visits during the intensive phase of treatment and 5.2 (SD: 1.8) if in the continuation phase of treatment. This is compared to 1.7 (SD: 0.5) during intensive – and 1.7 (SD: 0.2) during the continuation phase of treatment in the Labels arm of the study. Similarly, in the Pillbox study arm, patients had an average of 1.7 (SD: 0.2) visits during the intensive phase and 1.6 (SD: 0.2) during the continuation phase of treatment.

**Table 2.**
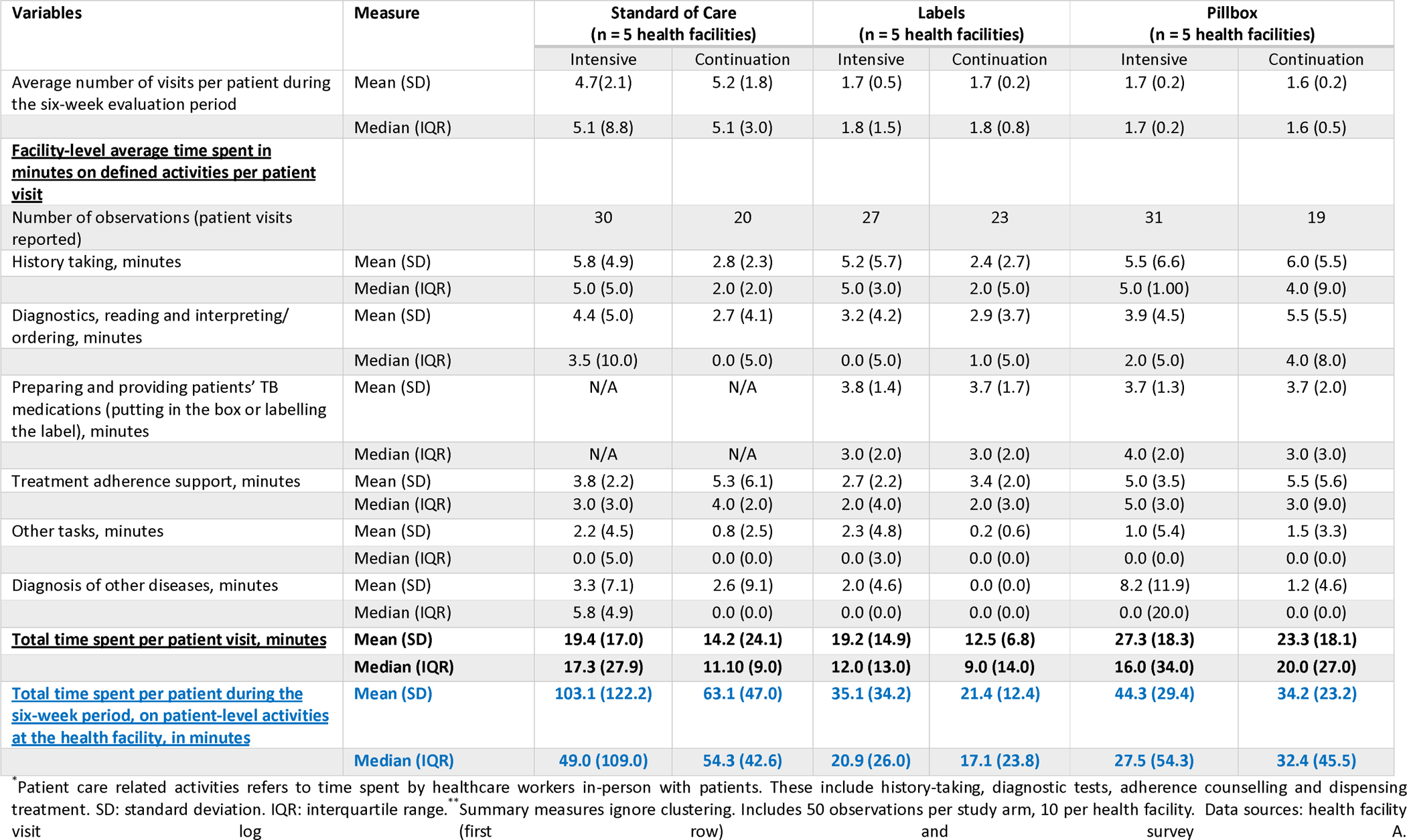
Facility-level average of patient-level observations. Health care provider time spent per patient per visit on direct patient care, by activity type ^*^ during the six-week evaluation period ^**^.

| Variables | Measure | Standard of Care<br>(n = 5 health facilities) |  | Labels<br>(n = 5 health facilities) |  | Pillbox<br>(n = 5 health facilities) |  |
| --- | --- | --- | --- | --- | --- | --- | --- |
|  |  | Intensive | Continuation | Intensive | Continuation | Intensive | Continuation |
| Average number of visits per patient during the six-week evaluation period | Mean (SD) | 4.7 (2.1) | 5.2 (1.8) | 1.7 (0.5) | 1.7 (0.2) | 1.7 (0.2) | 1.6 (0.2) |
|  | Median (IQR) | 5.1 (8.8) | 5.1 (3.0) | 1.8 (1.5) | 1.8 (0.8) | 1.7 (0.2) | 1.6 (0.5) |
| **<u>Facility-level average time spent in minutes on defined activities per patient visit</u>** |  |  |  |  |  |  |  |
| Number of observations (patient visits reported) |  | 30 | 20 | 27 | 23 | 31 | 19 |
| History taking, minutes | Mean (SD) | 5.8 (4.9) | 2.8 (2.3) | 5.2 (5.7) | 2.4 (2.7) | 5.5 (6.6) | 6.0 (5.5) |
|  | Median (IQR) | 5.0 (5.0) | 2.0 (2.0) | 5.0 (3.0) | 2.0 (5.0) | 5.0 (1.00) | 4.0 (9.0) |
| Diagnostics, reading and interpreting/ ordering, minutes | Mean (SD) | 4.4 (5.0) | 2.7 (4.1) | 3.2 (4.2) | 2.9 (3.7) | 3.9 (4.5) | 5.5 (5.5) |
|  | Median (IQR) | 3.5 (10.0) | 0.0 (5.0) | 0.0 (5.0) | 1.0 (5.0) | 2.0 (5.0) | 4.0 (8.0) |
| Preparing and providing patients' TB medications (putting in the box or labelling the label), minutes | Mean (SD) | N/A | N/A | 3.8 (1.4) | 3.7 (1.7) | 3.7 (1.3) | 3.7 (2.0) |
|  | Median (IQR) | N/A | N/A | 3.0 (2.0) | 3.0 (2.0) | 4.0 (2.0) | 3.0 (3.0) |
| Treatment adherence support, minutes | Mean (SD) | 3.8 (2.2) | 5.3 (6.1) | 2.7 (2.2) | 3.4 (2.0) | 5.0 (3.5) | 5.5 (5.6) |
|  | Median (IQR) | 3.0 (3.0) | 4.0 (2.0) | 2.0 (4.0) | 2.0 (3.0) | 5.0 (3.0) | 3.0 (9.0) |
| Other tasks, minutes | Mean (SD) | 2.2 (4.5) | 0.8 (2.5) | 2.3 (4.8) | 0.2 (0.6) | 1.0 (5.4) | 1.5 (3.3) |
|  | Median (IQR) | 0.0 (5.0) | 0.0 (0.0) | 0.0 (3.0) | 0.0 (0.0) | 0.0 (0.0) | 0.0 (0.0) |
| Diagnosis of other diseases, minutes | Mean (SD) | 3.3 (7.1) | 2.6 (9.1) | 2.0 (4.6) | 0.0 (0.0) | 8.2 (11.9) | 1.2 (4.6) |
|  | Median (IQR) | 5.8 (4.9) | 0.0 (0.0) | 0.0 (0.0) | 0.0 (0.0) | 0.0 (20.0) | 0.0 (0.0) |
| **<u>Total time spent per patient visit, minutes</u>** | Mean (SD) | **19.4 (17.0)** | **14.2 (24.1)** | **19.2 (14.9)** | **12.5 (6.8)** | **27.3 (18.3)** | **23.3 (18.1)** |
|  | Median (IQR) | **17.3 (27.9)** | **11.10 (9.0)** | **12.0 (13.0)** | **9.0 (14.0)** | **16.0 (34.0)** | **20.0 (27.0)** |
| **<u>Total time spent per patient during the six-week period, on patient-level activities at the health facility, in minutes</u>** | Mean (SD) | **103.1 (122.2)** | **63.1 (47.0)** | **35.1 (34.2)** | **21.4 (12.4)** | **44.3 (29.4)** | **34.2 (23.2)** |
|  | Median (IQR) | **49.0 (109.0)** | **54.3 (42.6)** | **20.9 (26.0)** | **17.1 (23.8)** | **27.5 (54.3)** | **32.4 (45.5)** |

\*Patient care related activities refers to time spent by healthcare workers in-person with patients. These include history-taking, diagnostic tests, adherence counselling and dispensing treatment. SD: standard deviation. IQR: interquartile range. \*\*Summary measures ignore clustering. Includes 50 observations per study arm, 10 per health facility. Data sources: health facility visit log (first row) and survey A.

Healthcare workers spend similar amounts of time on history taking between the three study arms for patients in the intensive phase of care but less time in the Labels and SoC arms if patients are in the continuation phase of treatment. A similar reduction is observed in time spent on diagnostics. For patients in the SoC arm of the study, there is a time saving of 3.7 minutes per patient visit compared to the Pillbox and Labels arms of the study because of time spent on preparing medication. Overall, healthcare workers were spending a mean of 27.3 (SD: 18.3) for those in the intensive phase and 23.3 (SD: 18.1) minutes per patient visit for those in the continuation phase if in the Pillbox arm of the study. This is compared to a mean of 19.2 (SD: 14.9) for intensive phase and 12.5 (SD: 6.8) for those in the Labels arms. Comparatively, healthcare workers were spending slightly less time with the patient per visit at 19.4 (17.0) minutes in the intensive phase and 14.2 (SD: 24.1) minutes in the continuation phase with those in the SoC arm (see Table 2). Our estimate of total time spent on patient-level activities in the SoC arm was highly right-skewed because of high variability between health facilities with two health facilities in the standard of care arm where daily visits were made to the health facility while in others there were only weekly or two-weekly visits.

Comparing the amount of time spent on above-service tasks or tasks when the patient is not at the clinic, we find that health care workers in the SoC arm facilities are spending an average of 121.0 (SD: 93.7) minutes or approximately two hours per week reviewing adherence records, compared to 95.0 (SD: 67.5) minutes in the Labels facilities and 45.3 (SD: 21.3) minutes in the Pillbox facilities (Table 3). This might be explained by the time used in SoC facilities to complete paper-based records documenting treatment adherence. In the Pillbox and Labels facilities, this function is replaced by the electronic adherence platform accessed by healthcare workers. In total for the six weeks, health care workers were spending 62.2 (SD: 86.0) hours in the standard of care arm, compared to 27.1 (SD: 23.7) hours in the Labels and 31.3 (25.8) hours in the Pillbox arms of the study.

**Table 3.** Facility-level observations. Human resource time spent on above-service activities and total time spent for all patients being seen over the six-week period per facility.

| Variables | Measure | Standard of Care<br>(n = 5 health<br>facilities) | Labels<br>(n = 5 health<br>facilities) | Pillbox<br>(n = 5 health<br>facilities) |
| --- | --- | --- | --- | --- |
| Average number of patients seen per facility during the six-week evaluation period. |  | 21.4 (7.6) | 16.6 (6.1) | 21.4 (4.4) |
| **<u>Facility-level average above-service time spent per week of the six-week evaluation period</u>** |  |  |  |  |
| Number of observations |  | 30 | 30 | 30 |
| Reviewing adherence records, minutes | Mean (SD) | 121 (93.7) | 95.0 (67.5) | 45.3 (21.3) |
|  | Median (IQR) | 90.0 (75.0) | 60.0 (60.0) | 45.0 (30.0) |
| Calling patients, minutes | Mean (SD) | 44.8 (39.1) | 80.0 (62.1) | 50.4 (33.8) |
|  | Median (IQR) | 45.0 (42.0) | 60.0 (90.0) | 60.0 (32.0) |
| Home visits, minutes | Mean (SD) | 2.0 (11.0) | 11.0 (39.0) | 26.0 (56.1) |
|  | Median (IQR) | 0.0 (0.0) | 0.0 (0.0) | 0.0 (0.0) |
| Administrative tasks, minutes | Mean (SD) | 225.4 (209.6) | 190.0 (213.5) | 221.0 (254.3) |
|  | Median (IQR) | 120.0 (300.0) | 120.0 (123.9) | 120.0 (180.0) |
| **<u>Total above-service time spent per week, in minutes</u>** | **Mean (SD)** | **393.2 (312.3)** | **376.9 (233.4)** | **342.7 (252.9)** |
|  | **Median (IQR)** | **240.0 (330.0)** | **320.1 (405.0)** | **270.0 (347.3)** |
| Number of observations |  | 5 | 5 | 5 |
| **<u>Total above-service plus direct patient care time spent per patient at the health facility during the six-week evaluation period in hours.</u>** | **Mean (SD)** | **62.2 (86.0)** | **27.1 (23.7)** | **31.3 (25.8)** |
|  | **Median (IQR)** | **37.6 (51.0)** | **17.9 (22.4)** | **19.1 (34.8)** |
| **<u>Human resource cost of tuberculosis care for the six-week period of evaluation. Average facility-level cost by study arm (USD).</u>** | **Mean (SD)** | **\$84.33 (43.88)** | **\$51.84 (32.94)** | **\$55.76 (30.23)** |
| | **Median (IQR)** | **\$85.00 (79.98)** | **\$44.55 (63.25)** | **\$55.26 (25.68)** |
| **<u>Average total human resource cost per patient (USD) during the six-week period of evaluation. Average of facilities by study arm.</u>** | **Mean (SD)** | **\$3.28 (2.07)** | **\$2.53 (1.82)** | **\$2.02 (1.07)** |
| | **Median (IQR)** | **\$2.57 (4.02)** | **\$1.87 (2.43)** | **\$1.87 (1.64)** |

Data sources: health facility visit log (first row) and survey B. With SD: standard deviation; IQR: interquartile range; USD: United States Dollars. Clustering was ignored in the calculation of the SDs per week. The average number of patients seen was estimated from the health facility visit log and the screening and enrolment log.

Table 4 shows the results of the comparative costing of staff time used to provide tuberculosis treatment support, comparing health facilities in the SoC to the Labels and Pillbox study arms. Overall, we find that the Labels and Pillbox arms are cost-saving per patient when compared the SoC though these findings are not statistically significant after adjusting for facility workload, and patients lost-to-follow-up (LTFU) with fairly wide confidence intervals (see Table 5). We found evidence of costs savings of $32.49 in the Labels and $28.57 in the Pillbox compared to the SoC health facilities, this was somewhat offset by training and support costs that were $2.89 and $3.88 higher in the Labels and Pillbox arms compared to the SoC arm of the study.

**Table 4.** Facility-level costs of human resources for providing tuberculosis treatment and adherence support for the six-week evaluation period.

|  | **Standard of Care**<br>**(n = 5 health facilities)** | **Labels**<br>**(n = 5 health facilities)** | **Pillbox**<br>**(n = 5 health facilities)** |
| --- | --- | --- | --- |
| **Human resources costs**<br>The cost of healthcare worker's time valued based on the facility specific salary costs (USD). Mean (SD). | \$84.33 (43.88) | \$51.84 (32.94) | \$55.76 (30.23) |
| **Training costs**<br>The cost of three rounds of training healthcare workers to implement the intervention, allocated to the six-week period (USD). Mean (SD). | \$0.12 (0.03) | \$0.30 (0.10) | \$0.23 (0.02) |
| **Implementation support costs.** Cost of providing support visits and calls to healthcare workers in implementing the intervention (USD). Mean (SD). | \$0.00 (0.00) | \$2.59 (0.23) | \$3.65 (1.10) |
| **Total facility-level human resource costs for tuberculosis care and treatment adherence support in 2023 USD** Mean (SD). | **\$84.45 (43.86)** | **\$54.73 (32.96)** | **\$59.64 (29.74)** |

Further details related to the cost analyses and cost assumptions are available in the appendix (see SS1). Implementation support costs includes the staff costs of calls and visits made by KNCV Ethiopia staff to health facilities in supporting the implementation of the interventions and distributing the Digital Adherence Technologies (DATs) within country.

**Table 5.**
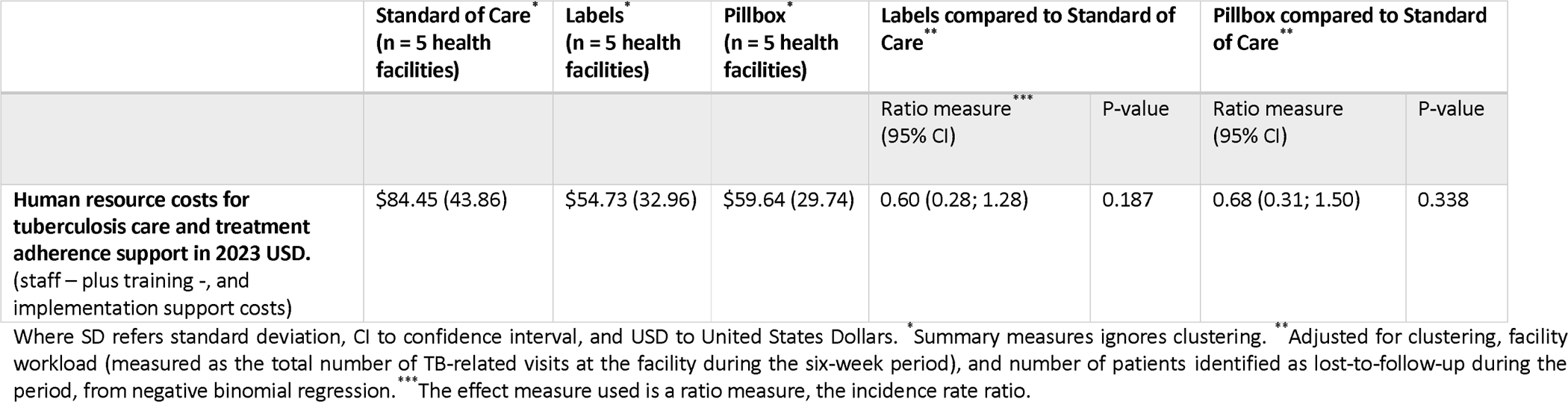
Effect of the implementation of Digital Adherence Technologies on the cost of human resource time use between study arms.

In a sensitivity analysis, where we varied key parameters to identify how the findings of our study may change if circumstances were different (see Figure 2), we found that increasing the cost of health care workers time (salary costs) would increase the cost savings from implementing DATs, while increased training and support costs would decrease these cost savings. Our findings were sensitive to assumptions related to the frequency of retraining required (see Figure 2). Our primary estimate includes the costs of three rounds of training on the use of DATs and associated guidelines. In our sensitivity analysis, assuming that retraining would be required every 4 months made the cost of DATs trend towards being cost neutral.

**Figure 2.**
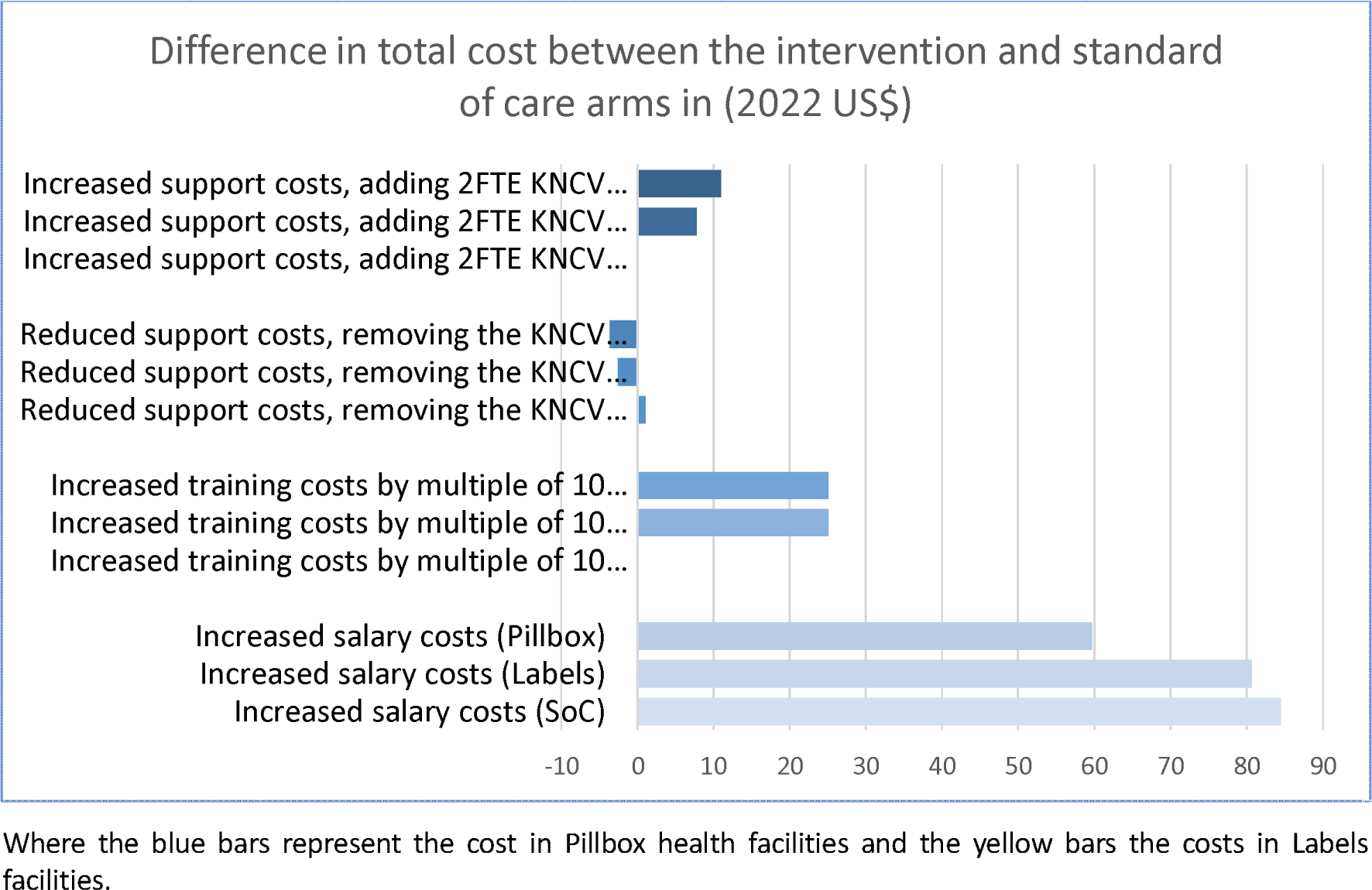
Sensitivity analyses.

## Discussion

We compared how health care workers spend their time supporting tuberculosis treatment adherence between standard of care and intervention health facilities where two types of digital adherence technologies were being implemented. We did not find a significant difference in time spent between study arms but found that healthcare workers were spending more time on in-person adherence counselling and less time on administrative tasks in the intervention arms compared to the SoC arm.

While we found that there were fewer patient visits in the Pillbox compared to the Labels and SoC facilities, healthcare workers were spending more time per patient during those in-person visits providing adherence support, especially in the intensive phase of treatment. Compared to the Labels arm, more intensive phase visits were reported in the SoC and Pillbox arms of the study, and as these visits take longer the total time estimates in these arms may be skewed compared to the Labels arm. Conversely, slightly more patients were identified as loss-to-follow up and requiring additional actions by health care workers during the evaluation period in the intervention compared to the SoC arms of the study possibly because HCWs had an easier way of identifying LTFU through the platform (see Table A in SS1). Patients identified as LTFU, suggests additional workload at the facility level as healthcare workers are calling and visiting patients prior to the numbers of LTFU being recorded.

Studies evaluating the cost and cost-effectiveness of digital adherence technologies for tuberculosis treatment reported to date have not directly measured the amount of time spent by healthcare workers on adherence support and compared this time between a SoC and intervention arm, however the cost of staff care have been included [10–12]. These studies found DATs to be more expensive than SoC and with mixed evidence of treatment effect. Nsengiyumva et al. estimated provider costs per person for 99DOTS (similar to our Labels arm) and Video Supported Treatment (VST) at $98 in Bangladesh (n = 719), $106 in the Philippines (n = 396) and $174 in Tanzania (n = 976) [10]. The study included data from a wide range of settings and used activities review and observation to collect data required for the cost analysis [10]. They concluded that labour cost offsets and/or economies of scale may yield savings when implementing 99DOTS relative to standard DOT. This assumes that DATs would save healthcare worker time, which is not what we found in our analysis. In Uganda, a step-wedged trial of medication sleeves found no evidence of improved treatment success during the intervention period in the ITT population but found improved outcomes in the PP population which was used to estimate cost-effectiveness. They estimated that the intervention cost US$355 (US$229; US$394) per treatment success based on the trial per protocol analysis that only includes those who successful completed treatment using the DAT [11][13]. A study conducted in Morocco, estimating the cost-effectiveness analysis of the pillbox for an observational cohort of MDR-TB patients. They used a Markov model to estimate costs from the provider perspective and found that the DAT compared to SoC was $434 per DALY averted [14]. These studies used activity logs to cost the use of DATs and did not compare the detailed time of human resource use for healthcare workers between study arms or between activities.

The shift from administrative tasks done when not with the patient to more time spent directly with patients including adherence counselling may explain some of the findings from DAT acceptability studies and some of the positive experiences of healthcare workers from qualitative surveys [15]. An acceptability study conducted as part of a step-wedged trial in Uganda showed that health care workers gave high acceptability scores of 99DOTS and patients there was a high level of agreement between patients that the DAT makes them feel more connected to healthcare workers [16].

In this study, we focused the cost analysis on the staff time required to implement the DATs. The full opportunity costs of the implementation of DATs including the technology costs, medication, diagnostics used for monitoring, and subsequent treatment episodes will be assessed in a separate cost-utility analysis [8]. Some of the limitations of our study include that it was based on healthcare workers self-completing a time-and-motion survey retrospectively, at the end of each patient seen or at the end of the week for above-service tasks. Healthcare workers self-completing the survey, may be biased due to social desirability bias with healthcare workers reporting estimates based on what they think the implementing organisation is desiring and potential recall bias. Another approach that we could have taken would have been if we observed healthcare workers in their tasks, noting down the time. However, direct observation may impact on service provision and would potentially underestimate above service level tasks completed when the patient (and observer) is not at the health facility. We have mitigated this potential underestimation by collecting additional data on time spent on above-service level activities, training, and support. Furthermore, our approach of estimating costs using a bottom-up approach underestimates the full cost of employing staff to provide the service in health facilities with fewer patients seen. It may be that in more rural health facilities, where there are fewer patients in need of tuberculosis care, a healthcare worker would be employed to provide such care but would not be seeing tuberculosis patients for the entire time. Nevertheless, one may argue that such healthcare workers may then be working elsewhere in the health facility where needed during busy times. The cost per patient reported in this paper should be interpreted with caution. Traditionally, cost per patient estimates would include costs incurred for the entire treatment episode. In this analysis, we were comparing activities between study arms for a six-week period only. This cost per patient estimates therefore only include provider costs incurred due to activities during the six-week period of analysis and not for the entire treatment episode (∼6 months) limiting comparability of this estimate. Despite these limitations, our study is the first of its kind to explicitly assess differences in staff time spent on tuberculosis treatment adherence support between SoC and DAT health facilities.

Our findings have implications for the planning of human resources in Ethiopia and the roll-out of tuberculosis treatment adherence support. DATs do not reduce time spent by healthcare workers on patient care. However, they result in fewer visits to health facilities, with healthcare workers spending more time on in-person patient care. We did observe a reduction in adherence monitoring work in facilities implementing DAT, however this was offset by time spent on phone calls to patients and home visits. Further work is needed to understand the impact that digital adherence monitoring may have on tuberculosis treatment outcomes, and the cost-effectiveness of DATs.

## Acknowledgements

This study is part of the ASCENT consortium, which is a partnership between KNCV Tuberculosis Foundation, The Aurum Institute, The London School of Hygiene & Tropical Medicine (LSHTM), the governments of the implementing countries and PATH. The project is funded by Unitaid. This work would not have been possible without the health facility staff who implemented the intervention, the support of the country national tuberculosis program, the oversight from the ASCENT Technical Advisory Group and the study participants who participated in the study.

## Data availability

Facility-level data analysed in this study are part of the ASCENT Consortium Trials data and will be made available with the publication of the paper describing the ASCENT-Ethiopia within-trial cost-effectiveness analysis. A description of the ASCENT-Ethiopia 6-week provider costs dataset, the survey materials, plus the code required to reproduce the analysis may be found at https://doi.org/10.17037/DATA.00003801. Following publication of the trial cost-effectiveness results, the minimal dataset will be available for non-commercial use upon request, after signing a data sharing agreement, to studies approved by an ethics committee. Any publications arising from the shared data must acknowledge the investigators who collected the data, the institutions involved, and the funding sources by citing the data record and including the standard acknowledgement statement provided.

## Author contributions

Conceptualisation: NF, MB, MS, AWT, KLF; Data curation: MS, MB, DG; NF; Formal analysis: MB, NF; Funding acquisition: KvK, KLF; Methodology: MB, MS, NF, KLF; Resources: TA, AB, JvR, KvK; Supervision: AWT, TA, AB, KLF, NF, DJ; Writing – Original Draft Preparation: MB, NF; Writing – Review and Editing: MB, MS, DG, TA, AB, CFM, JvR, KvK, DJ, KLF, AWT, NF.

