## Supplementary material for "Effect of implementing digital adherence technologies on the use of health care providers’ time and the human resource cost of tuberculosis treatment adherence support in Ethiopia": ss1 text

### 1. Producing datasets for analysis

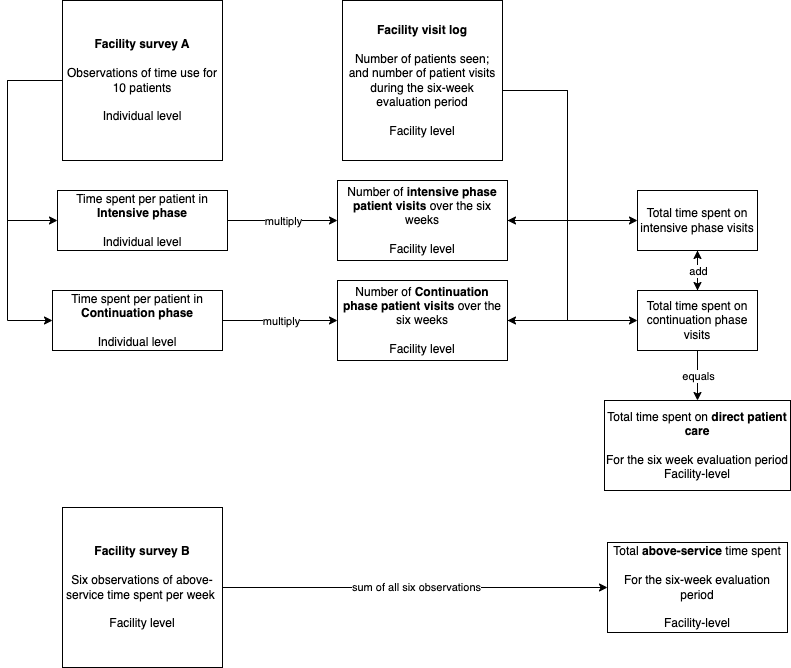

Figure A. Flow chart summarising the production of datasets for analysis.

### 2. Facility summaries

Data of the timing of the survey period, number of missed doses and number of patients reported lost-to-follow-up during the period is summarised in Table S1.

Table A. Survey period and lost-to-follow-up data per facility.

| FacilityID | StudyArm | Start date | End date | Missed doses | Loss to follow-up |
| --- | --- | --- | --- | --- | --- |
| A | Pillbox | 04-10-21 | 14-11-21 | 0 | 1 |
| B | Labels | 08-04-22 | 17-05-22 | 0 | 1 |
| C | Pillbox | 06-05-22 | 15-06-22 | 0 | 0 |
| D | Pillbox | 08-11-21 | 21-12-21 | 0 | 0 |
| E | SOC | 07-02-22 | 18-03-22 | NA | 0 |
| F | Labels | 14-02-22 | 27-03-22 | 0 | 0 |
| G | SOC | 12-03-22 | 23-04-22 | NA | 1 |
| H | SOC | 09-01-22 | 24-02-22 | NA | 0 |
| I | Labels | 21-03-22 | 03-05-22 | 0 | 1 |
| J | Pillbox | 01-11-21 | 10-12-21 | 0 | 0 |
| K | Labels | 25-10-21 | 05-12-21 | 1 | 0 |
| L | SOC | 28-02-22 | 10-04-22 | NA | 0 |
| M | SOC | 24-01-22 | 09-04-22 | NA | 0 |
| N | Labels | 12-04-22 | 22-05-22 | 0 | 0 |
| O | Pillbox | 16-05-22 | 26-06-22 | 0 | 0 |

### 3. Cost analysis

We estimated three types of human resources costs incurred for the duration of the six-week period of evaluation, firstly the cost of the TB focal person based in the health facilities’ time, secondly the cost of training TB focal people from each health facility in implementing the interventions and thirdly the implementation support costs incurred by the implementing partner (KNCV-Ethiopia) in providing telephonic supports and physical visits to the health facilities.

### 4. Additional results

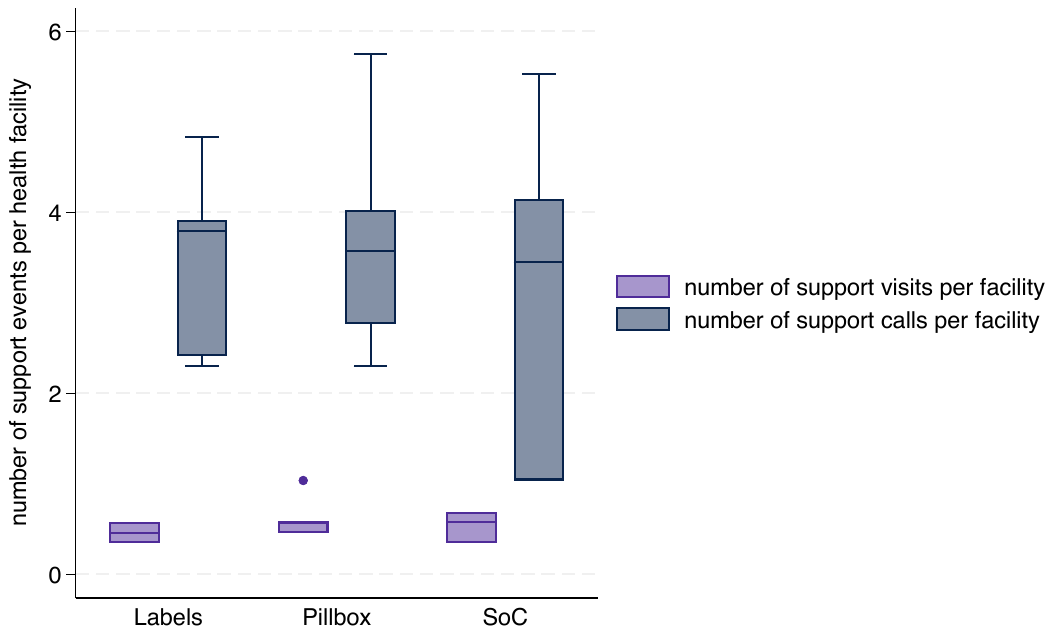

Figure B. Number of support events (visits and calls) per facility sampled during the six-week evaluation period.

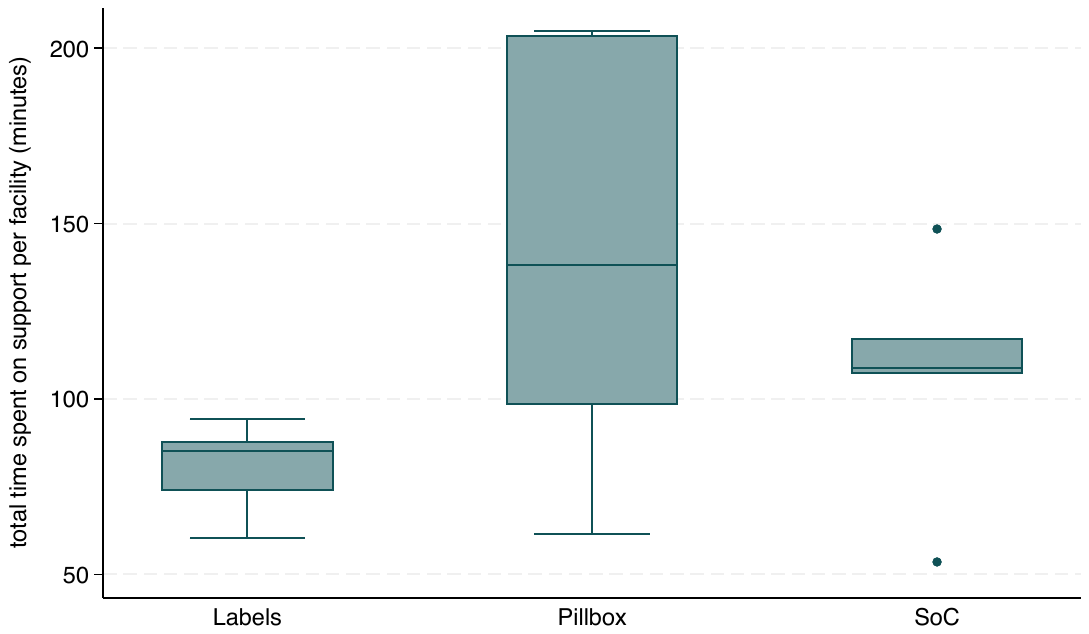

Figure C. Total time spent on support per facility sampled during the six-week evaluation period.
